# Perceptions and acceptance of COVID-19 vaccine among pregnant and lactating women in Singapore: A cross-sectional study

**DOI:** 10.1101/2021.06.29.21259741

**Authors:** Pooja A Jayagobi, Chengsi Ong, Yeo Kee Thai, Caleb CW Lim, Seet Meei Jiun, Kwek Lee Koon, Ku Chee Wai, Jerry KY Chan, Manisha Mathur, Chua Mei Chien

## Abstract

**Introduction:** Vaccination is critical in controlling the coronavirus disease 19 (COVID-19) pandemic. However, vaccine perception and acceptance among pregnant and lactating women is unknown in Singapore. We aimed to determine the acceptance of COVID-19 vaccination among these two groups of women in Singapore, and factors associated with vaccine acceptance.

**Methods:** We conducted an anonymous, online survey on the perception and acceptance of the COVID-19 vaccine in pregnant and lactating women at a tertiary hospital in Singapore from 1^st^ March to 31^st^ May 2021. Information on demographics and knowledge were collected, and these factors were assessed for their relationship with vaccine acceptance.

**Results:** A total of 201 pregnant and 207 lactating women participated. Vaccine acceptance rates in pregnant and lactating women were 30.3% and 16.9% respectively. Pregnant women who were unsure or unwilling to take the vaccine cited concerns about safety of the vaccine during pregnancy (92.9%), while lactating women were concerned about potential long-term negative effects on the breastfeeding child (75.6%). Other factors significantly associated with vaccine acceptance included a lower monthly household income or education level, appropriate knowledge regarding vaccine mechanism and higher perceived maternal risk of COVID-19. Most pregnant (70.0%) and lactating women (83.7%) were willing to take the vaccine only when more safety data during pregnancy and breastfeeding were available.

**Conclusions:** COVID-19 vaccine acceptance was low among pregnant and lactating women in Singapore. Addressing safety concerns when more data is available and education on mechanism of vaccine action will likely improve acceptance among these women.

## Introduction

In this current coronavirus disease 2019 (COVID-19) pandemic, vaccination remains a critical strategy to curbing infections and reducing disease severity. Current approved COVID-19 vaccines in Singapore (Pfizer-BioNtech and the Moderna COVID-19 mRNA vaccines) have been found to be safe and efficacious in preventing severe disease.^(1,2)^ As these initial vaccine trials did not include pregnant and lactating women, Singapore’s Ministry of Health (MOH) initially cautioned against vaccination in pregnant women, and recommended lactating women to stop breastfeeding for 5 to 7 days following vaccination.^(3,4)^ These were in contrast to recommendations from the Society for Maternal-Fetal Medicine (SMFM) and American College of Obstetricians and Gynecologists (ACOG) that have maintained that the vaccine should be offered to pregnant and lactating women based on their risk, and that the mRNA-based vaccines are thought to pose a low risk to the fetus as the mRNA is expected to degrade in circulation.^(5,6)^ The World Health Organization and the Academy of Breastfeeding Medicine recommend that lactating women should continue to breastfeed post vaccination as it is unlikely for mRNA-based vaccines to be transmitted via breast milk.^(7,8)^

With the emerging local and international clinical reports on the safety and efficacy of mRNA-based COVID-19 vaccines, MOH, Singapore approved the use of these vaccines for pregnant women and recommended lactating women to continue breastfeeding after vaccination on 31 May 2021. Pregnant women are a vulnerable group in this pandemic, with an increased risk for severe disease and adverse outcomes if infected, including preterm birth, venous thromboembolism, severe respiratory complications requiring invasive ventilation.^(9-12)^ Lactating mothers could also potentially infect the infant postnatally via droplet infection.^(13,14)^ Despite the benefits of vaccination to prevent maternal and fetal complications, studies have demonstrated varying rates of vaccine acceptance in pregnant and lactating women.^(15,16)^ Reported factors affecting acceptance included confidence in the safety and efficacy of the vaccine, level of trust in public health agencies, perceived lack of research and fear of harming the fetus.^(15,16)^

We undertook this survey to evaluate the perceptions and factors affecting acceptance of the COVID-19 vaccine among pregnant and lactating women in a tertiary care centre in Singapore. We propose that the results will inform healthcare providers and policymakers on reasons for vaccine refusal, which can help develop strategies to improve vaccine uptake in these groups of women.

## Methods

This was a cross-sectional, web-based survey conducted at a tertiary maternal and child hospital in Singapore. Pregnant and lactating women over 21 years of age, attending outpatient clinics or admitted to the hospital were invited to participate in an anonymous, online survey from 1^st^ March to 31^st^ May 2021. Participation was voluntary and no incentive was offered for participation. No personally identifiable data was collected. The survey was advertised at outpatient clinics, inpatient nurseries and obstetric wards, and among the hospital’s healthcare workers via posters and flyers. Ethics approval was obtained from the Singhealth Institutional Review Board (CIRB Ref No.: 2020/2648) with a waiver for informed consent.

Separate surveys were designed for lactating mothers and for pregnant women using input from other published studies and healthcare professionals.^(17,18)^ The surveys were developed and administered in English, and consisted of questions on demographics, vaccine acceptance, general health status, COVID-19 experience, knowledge regarding the COVID-19 vaccine, and participant perceptions and concerns regarding the COVID-19 vaccine. The full survey consisted of 22 questions for pregnant women and 28 questions for lactating mothers. Responses to all the questions were in multiple-choice format. Perceptions of COVID-19 vaccine were rated on a 5-point Likert Scale.

The surveys were hosted on an online self-service form builder which stores responses in an encrypted format and accessible only to the creator via a special code (Form.gov.sg, GovTech, Singapore). Entry into the survey was via a QR code printed on the invitation flyer given to the respondents. All the questions were mandatory and displayed in running order on the respondent’s screen. Respondents were prompted to complete the outstanding questions before submitting the form, and were unable to save responses or change their answers after submission.

All submitted surveys were included in the analysis of this study. Vaccine acceptance was defined as a response of “strongly agree” or “agree” to take the vaccine if offered, while vaccine non-acceptance was defined as a response of “unsure”, “disagree” or “strongly disagree”. Responses to knowledge questions were categorized into correct and wrong responses, while responses to perception questions were organized into similar categories (e.g., “strongly agree” and “agree” were taken to be one category).

Univariate logistic regression analysis was used to determine demographics, vaccine knowledge and vaccine safety perceptions factors associated with vaccine acceptance in pregnant and lactating women separately. Factors most strongly associated with vaccine acceptance in the univariate analyses were entered sequentially into a multivariable forward stepwise regression analysis with significance levels to enter/stay of 0.10/0.05. Model goodness-of-fit was also assessed using the Akaike information criterion (AIC). All data was analyzed using SPSS version 19.0 (IBM Corp, Armonk, USA). The Checklist for Reporting Results of Internet E-Surveys (CHERRIES) was used in the reporting of this study.^(19)^

## Results

A total of 201 pregnant women and 207 lactating women participated in the survey (**Table I)**. Majority of the respondents in both pregnant and lactating groups were 21-34 years old (77.6% and 66.7% respectively), Singaporean citizens (79.1% and 73.9%), of Chinese ethnicity (48.3% and 52.7%) and without pre-existing medical conditions (97% and 92.7%)

**Table I.**
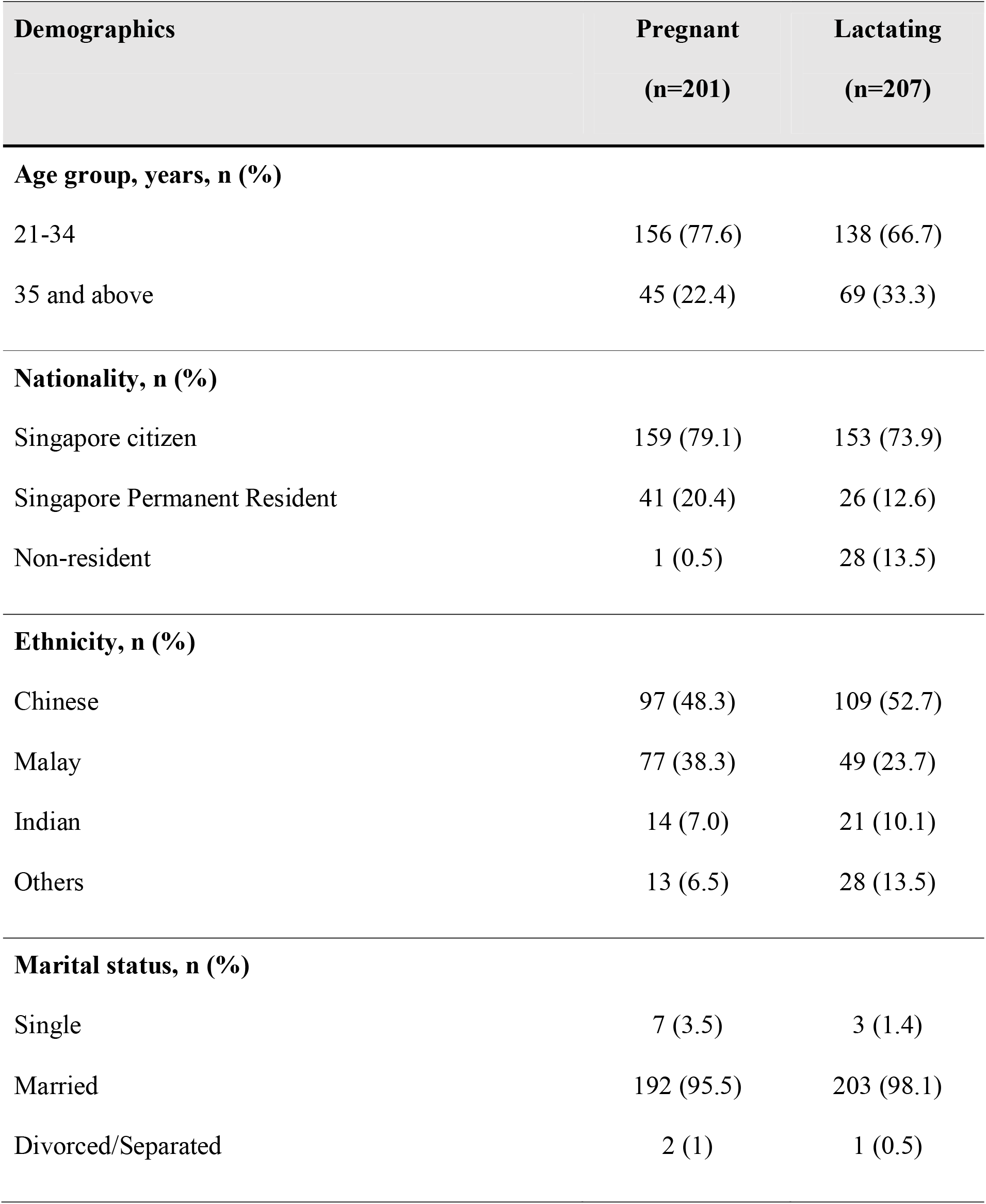
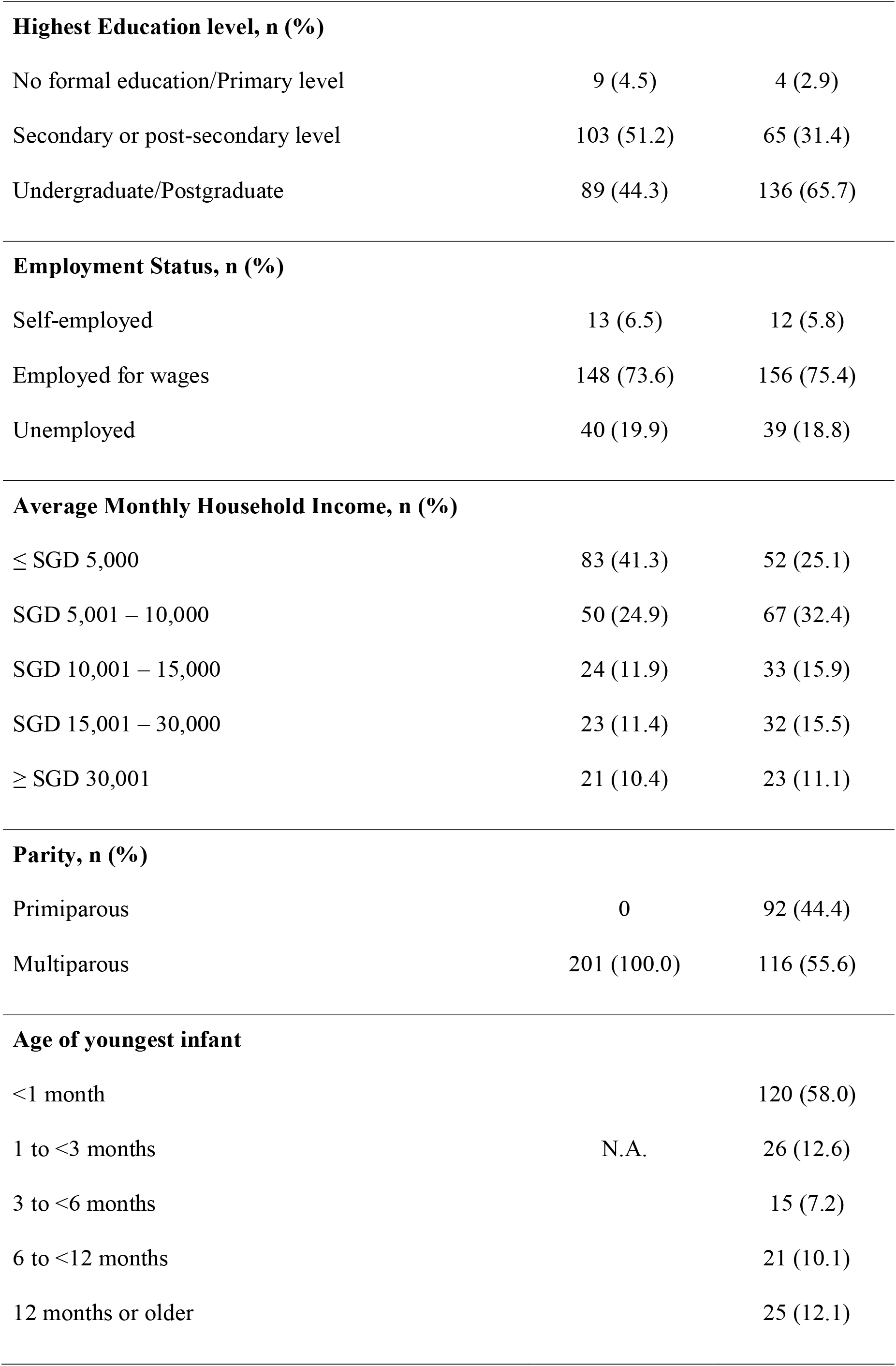
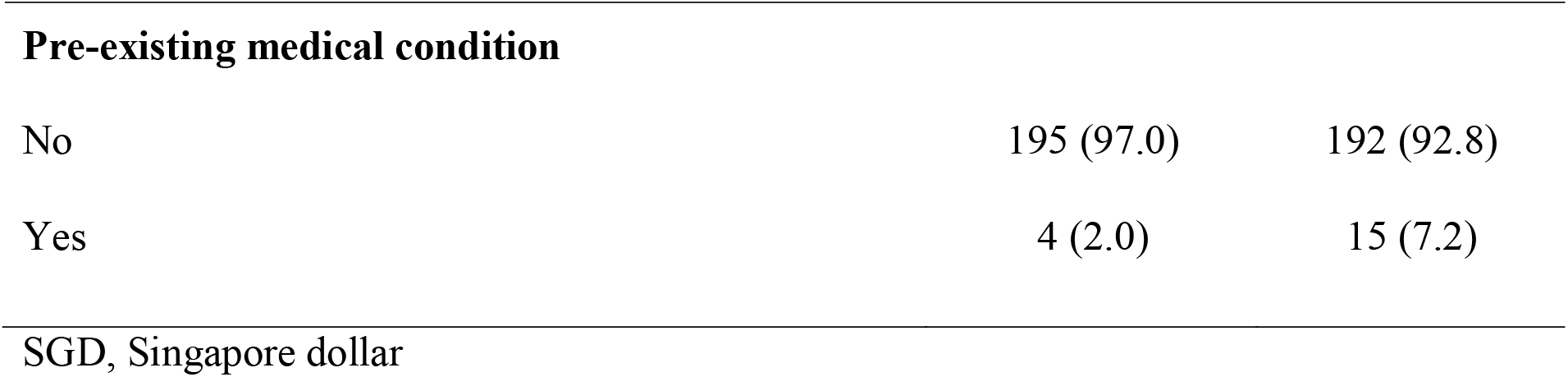
Participant characteristics of pregnant and lactating women.

### Pregnant women

The majority of pregnant women were aware of the mRNA vaccine’s mode of action (69.6%), however only 32.9% of pregnant women reported to be aware that the vaccine was efficacious in reducing risk of COVID-19 in pregnant women (**Table II**). Only 61/201 (30.3%) were agreeable to take the COVID-19 vaccine if it was offered to them, while the remainder were unsure (n=91, 45.3%) or unwilling (n=49, 24.4%). Of those who were unsure or unwilling to take the vaccine 92.9% (109/140) had doubts about the safety of the vaccine, 78.6% (110/140) were concerned about the side effects they would experience from the vaccine and 92.1% (129/140) were worried about the unknown short and long-term effects of the vaccine on the pregnancy and unborn child. Most in this group reported that they would take the vaccine only if there was more data available on its safety during pregnancy (119/140, 70.0%).

**Table II.**
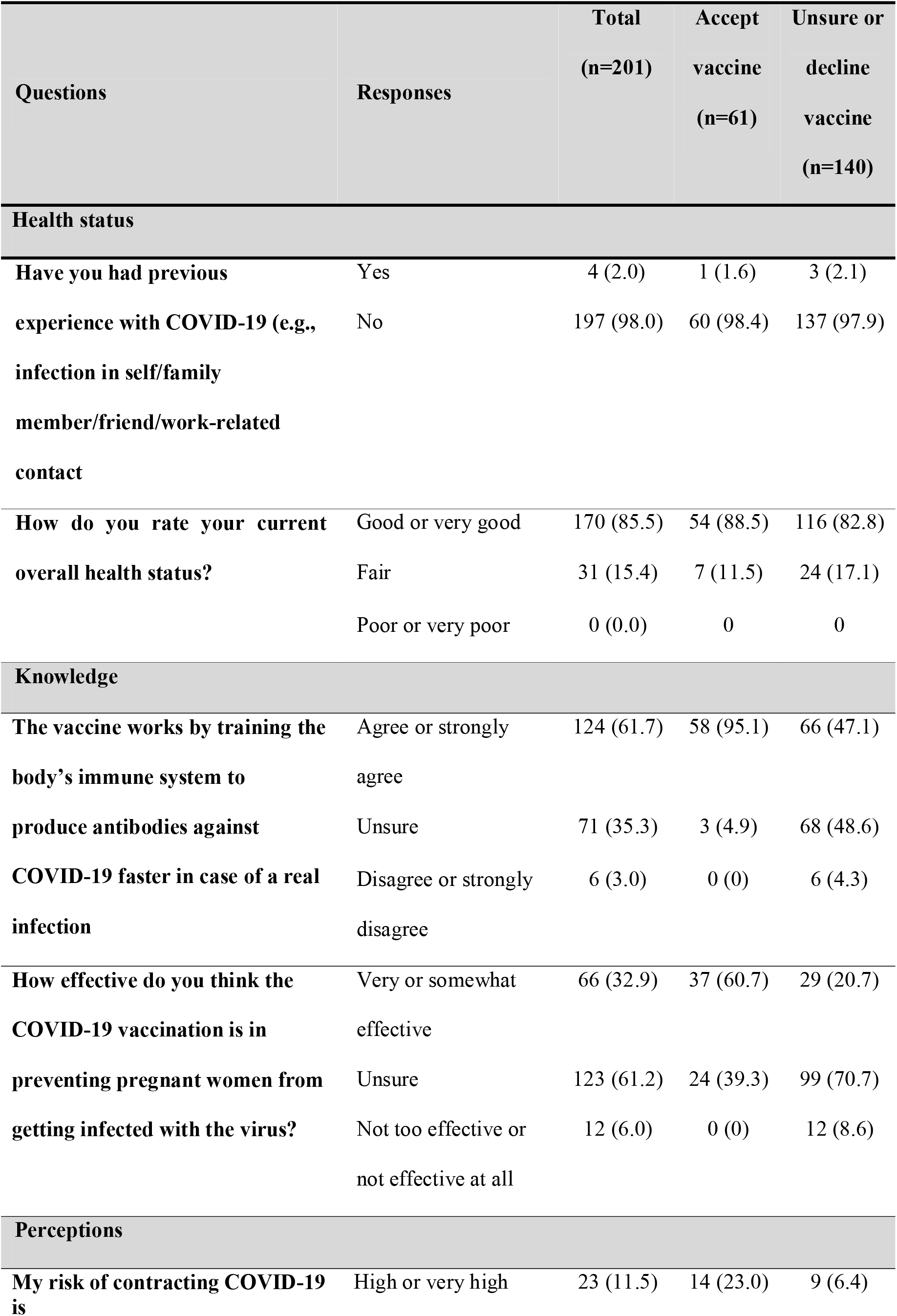
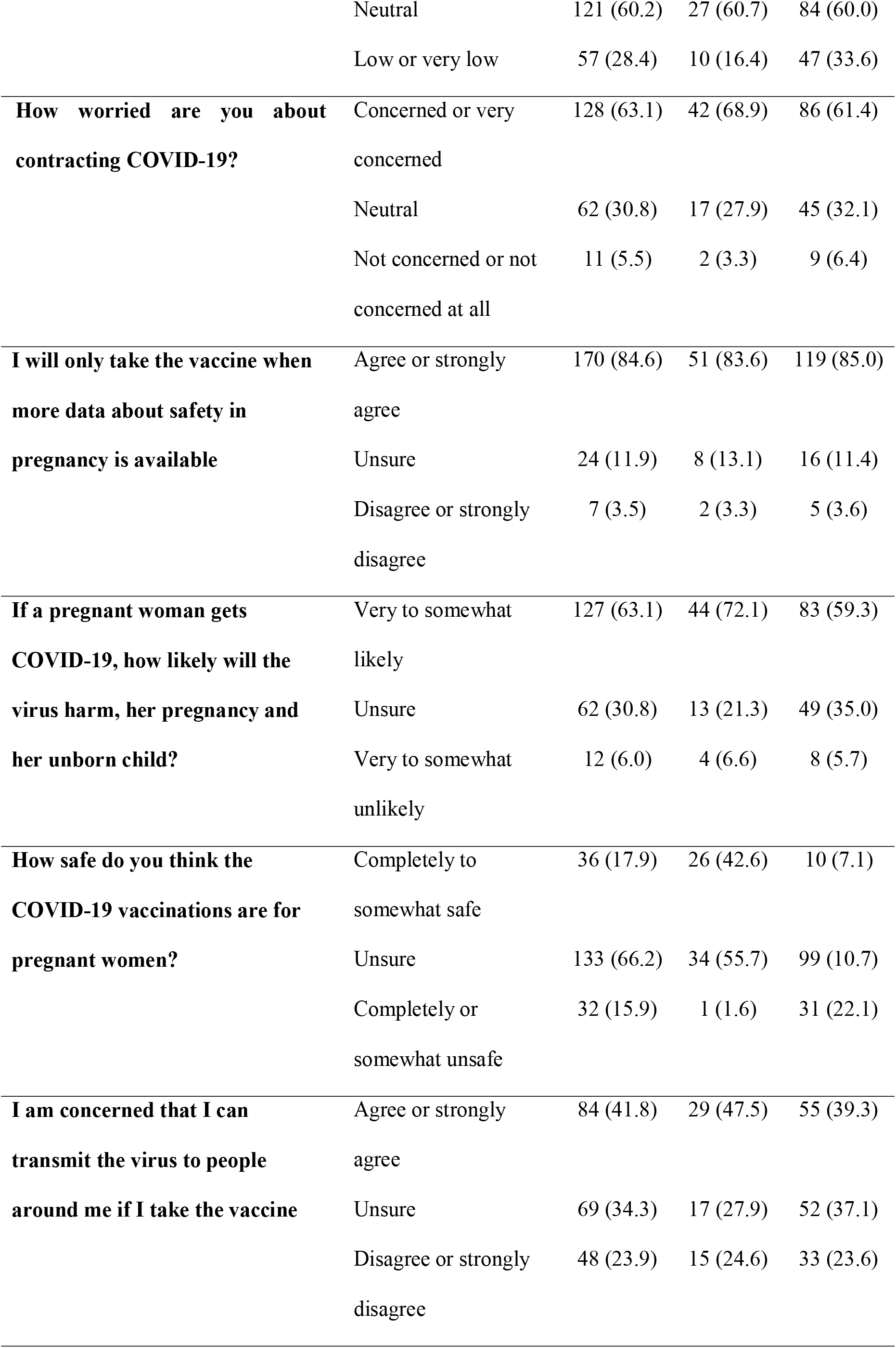
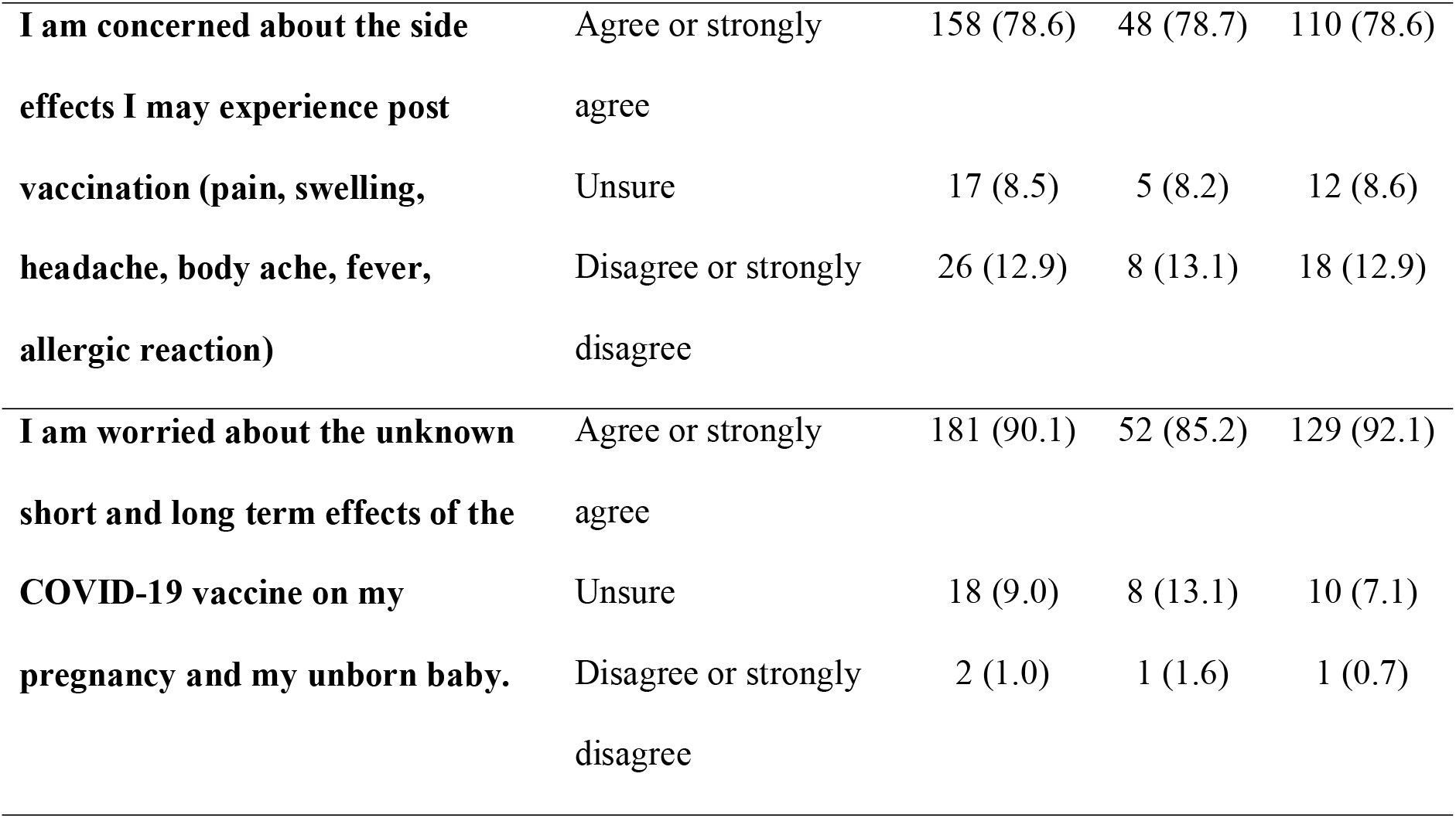
Survey responses for pregnant women.

| Questions | Responses | Total<br>(n=201) | Accept<br>vaccine<br>(n=61) | Unsure or<br>decline<br>vaccine<br>(n=140) |
| --- | --- | --- | --- | --- |
| **Health status** |  |  |  |  |
| Have you had previous experience with COVID-19 (e.g., infection in self/family member/friend/work-related contact) | Yes | 4 (2.0) | 1 (1.6) | 3 (2.1) |
|  | No | 197 (98.0) | 60 (98.4) | 137 (97.9) |
| How do you rate your current overall health status? | Good or very good | 170 (85.5) | 54 (88.5) | 116 (82.8) |
|  | Fair | 31 (15.4) | 7 (11.5) | 24 (17.1) |
|  | Poor or very poor | 0 (0.0) | 0 | 0 |
| **Knowledge** |  |  |  |  |
| The vaccine works by training the body's immune system to produce antibodies against COVID-19 faster in case of a real infection | Agree or strongly agree | 124 (61.7) | 58 (95.1) | 66 (47.1) |
|  | Unsure | 71 (35.3) | 3 (4.9) | 68 (48.6) |
|  | Disagree or strongly disagree | 6 (3.0) | 0 (0) | 6 (4.3) |
| How effective do you think the COVID-19 vaccination is in preventing pregnant women from getting infected with the virus? | Very or somewhat effective | 66 (32.9) | 37 (60.7) | 29 (20.7) |
|  | Unsure | 123 (61.2) | 24 (39.3) | 99 (70.7) |
|  | Not too effective or not effective at all | 12 (6.0) | 0 (0) | 12 (8.6) |
| **Perceptions** |  |  |  |  |
| My risk of contracting COVID-19 is | High or very high | 23 (11.5) | 14 (23.0) | 9 (6.4) |
|  | Neutral | 121 (60.2) | 27 (60.7) | 84 (60.0) |
|  | Low or very low | 57 (28.4) | 10 (16.4) | 47 (33.6) |
| **How worried are you about contracting COVID-19?** | Concerned or very concerned | 128 (63.1) | 42 (68.9) | 86 (61.4) |
|  | Neutral | 62 (30.8) | 17 (27.9) | 45 (32.1) |
|  | Not concerned or not concerned at all | 11 (5.5) | 2 (3.3) | 9 (6.4) |
| **I will only take the vaccine when more data about safety in pregnancy is available** | Agree or strongly agree | 170 (84.6) | 51 (83.6) | 119 (85.0) |
|  | Unsure | 24 (11.9) | 8 (13.1) | 16 (11.4) |
|  | Disagree or strongly disagree | 7 (3.5) | 2 (3.3) | 5 (3.6) |
| **If a pregnant woman gets COVID-19, how likely will the virus harm, her pregnancy and her unborn child?** | Very to somewhat likely | 127 (63.1) | 44 (72.1) | 83 (59.3) |
|  | Unsure | 62 (30.8) | 13 (21.3) | 49 (35.0) |
|  | Very to somewhat unlikely | 12 (6.0) | 4 (6.6) | 8 (5.7) |
| **How safe do you think the COVID-19 vaccinations are for pregnant women?** | Completely to somewhat safe | 36 (17.9) | 26 (42.6) | 10 (7.1) |
|  | Unsure | 133 (66.2) | 34 (55.7) | 99 (10.7) |
|  | Completely or somewhat unsafe | 32 (15.9) | 1 (1.6) | 31 (22.1) |
| **I am concerned that I can transmit the virus to people around me if I take the vaccine** | Agree or strongly agree | 84 (41.8) | 29 (47.5) | 55 (39.3) |
|  | Unsure | 69 (34.3) | 17 (27.9) | 52 (37.1) |
|  | Disagree or strongly disagree | 48 (23.9) | 15 (24.6) | 33 (23.6) |
| **I am concerned about the side effects I may experience post vaccination (pain, swelling, headache, body ache, fever, allergic reaction)** | Agree or strongly agree | 158 (78.6) | 48 (78.7) | 110 (78.6) |
|  | Unsure | 17 (8.5) | 5 (8.2) | 12 (8.6) |
|  | Disagree or strongly disagree | 26 (12.9) | 8 (13.1) | 18 (12.9) |
| **I am worried about the unknown short and long term effects of the COVID-19 vaccine on my pregnancy and my unborn baby.** | Agree or strongly agree | 181 (90.1) | 52 (85.2) | 129 (92.1) |
|  | Unsure | 18 (9.0) | 8 (13.1) | 10 (7.1) |
|  | Disagree or strongly disagree | 2 (1.0) | 1 (1.6) | 1 (0.7) |

Regression analysis demonstrated that factors associated with vaccine acceptance in pregnant women were a lower monthly household income, appropriate knowledge regarding vaccine mechanism, higher maternal perceived risk of COVID-19, and perceived general safety of the vaccine during pregnancy (**Table III**).

**Table III.** Multivariate regression analysis for vaccine acceptance in pregnant women.

| **Factors** | **Univariate**<br>**Odds ratio (95%**<br>**confidence interval)** | **p** |
| --- | --- | --- |
| **Monthly household income, SGD** |  |  |
| 5000 below | Ref | N.A. |
| 5001 to 15000 | 0.39 (0.16, 0.94) | 0.04 |
| Above 15000 | 0.23 (0.07, 0.78) | 0.02 |
| **Knowledge about mechanism of action** |  |  |
| Unsure/Disagree or strongly disagree | Ref | N.A. |
| Agree or strongly agree | 18.52 (5.05, 68.00) | <0.001 |
| **Maternal perceived risk of COVID** |  |  |
| Low or very low | Ref | N.A. |
| Neutral | 2.98 (1.13, 7.86) | 0.03 |
| High or very high | 6.25 (1.59, 24.56) | 0.009 |
| **Perceived general safety of vaccine** |  |  |
| Somewhat or completely unsafe | Ref | N.A. |
| Unsure | 5.23 (0.61, 44.96) | 0.13 |
| Somewhat or completely safe | 38.55 (3.93, 377.82) | 0.002 |
| SGD, Singapore dollar. |  |  |

### Lactating women

Up to 61.7% of lactating women were aware of the vaccine mode of action, and 72.9% were aware of its efficacy in reducing COVID-19 risk (**Table IV**). Among lactating women, 35 (16.9%) were agreeable to take the vaccine, while 129 (62.3%) were unsure, and 43 (20.8%) were unwilling.

**Table IV.**
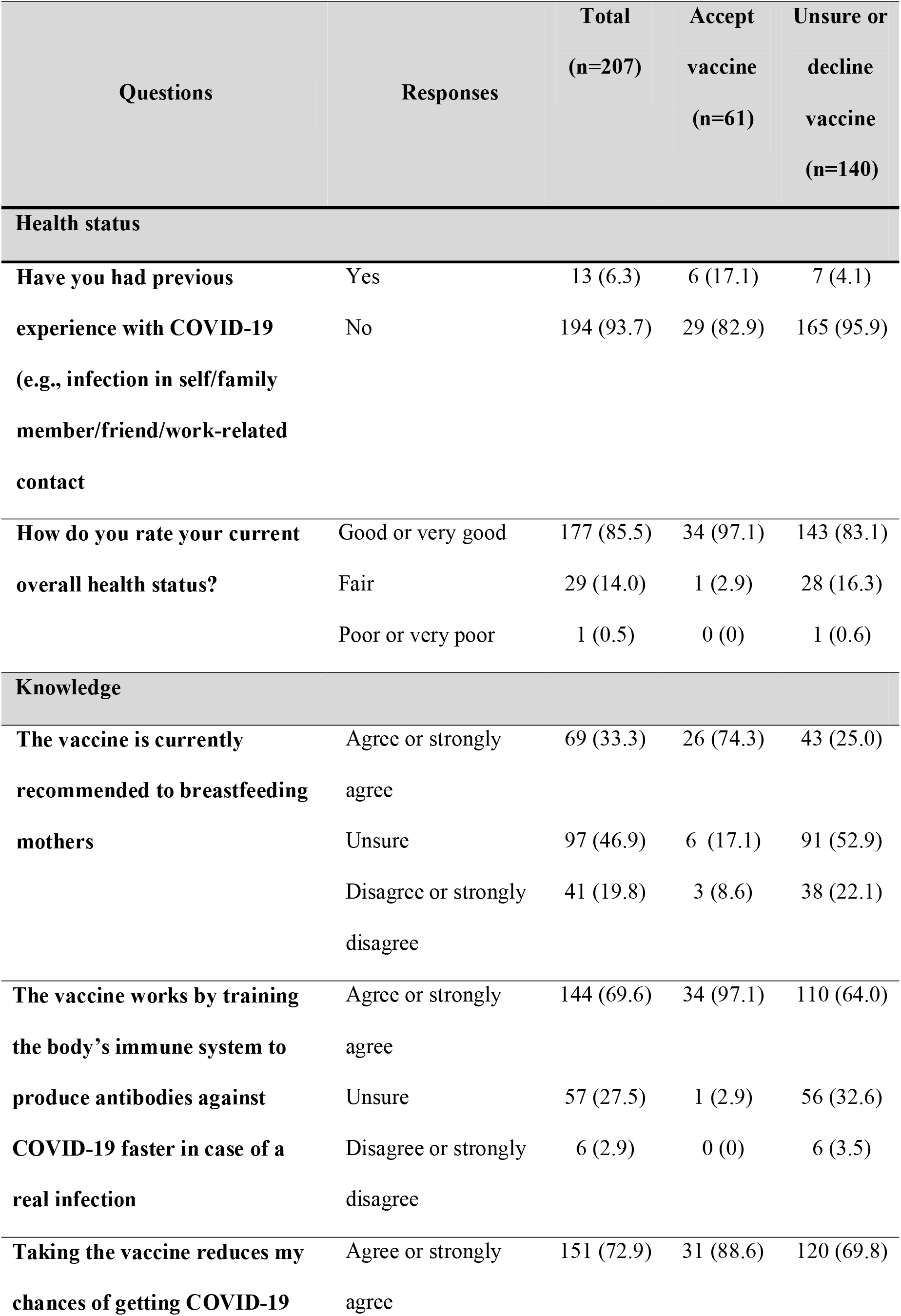
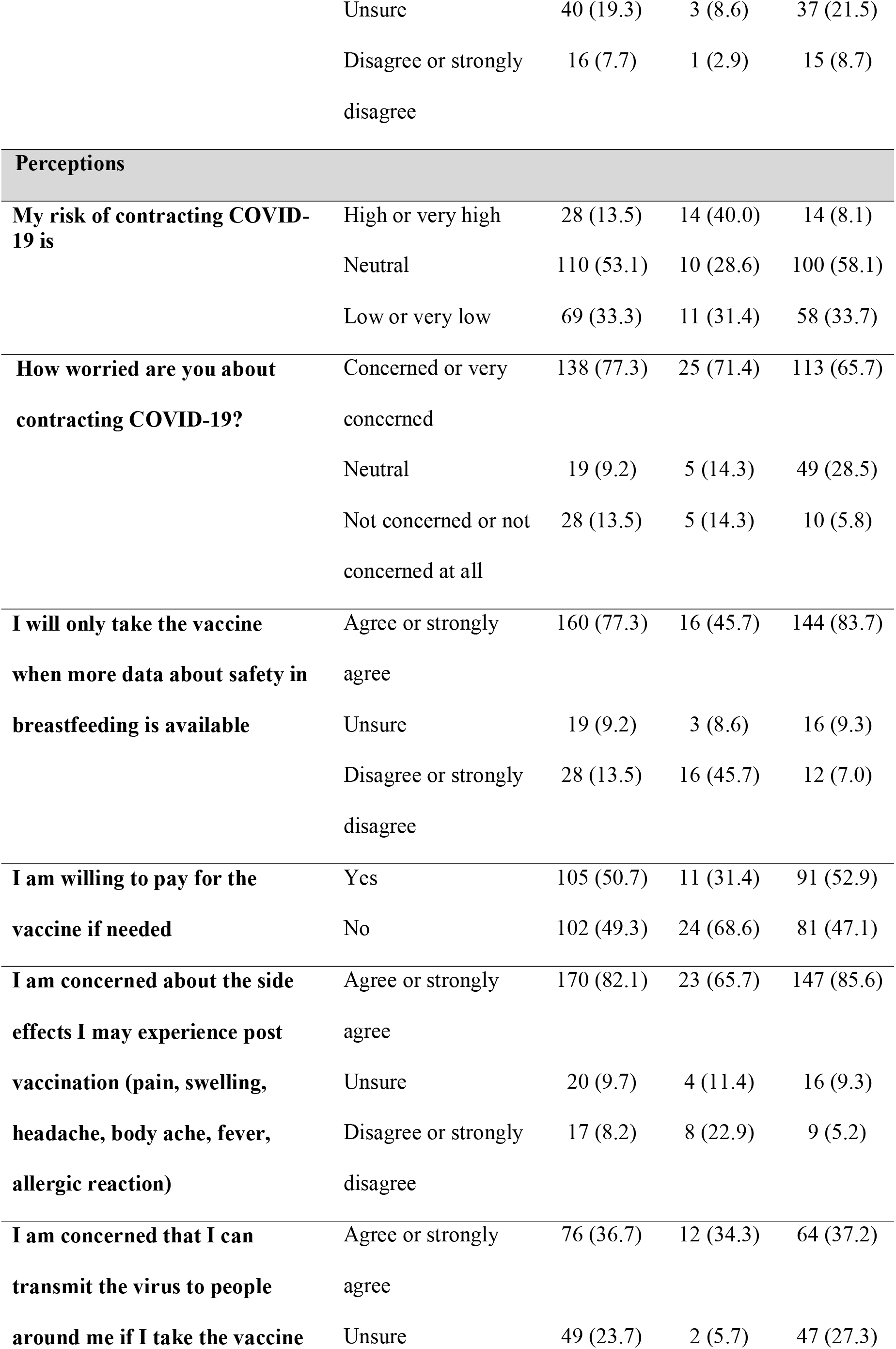
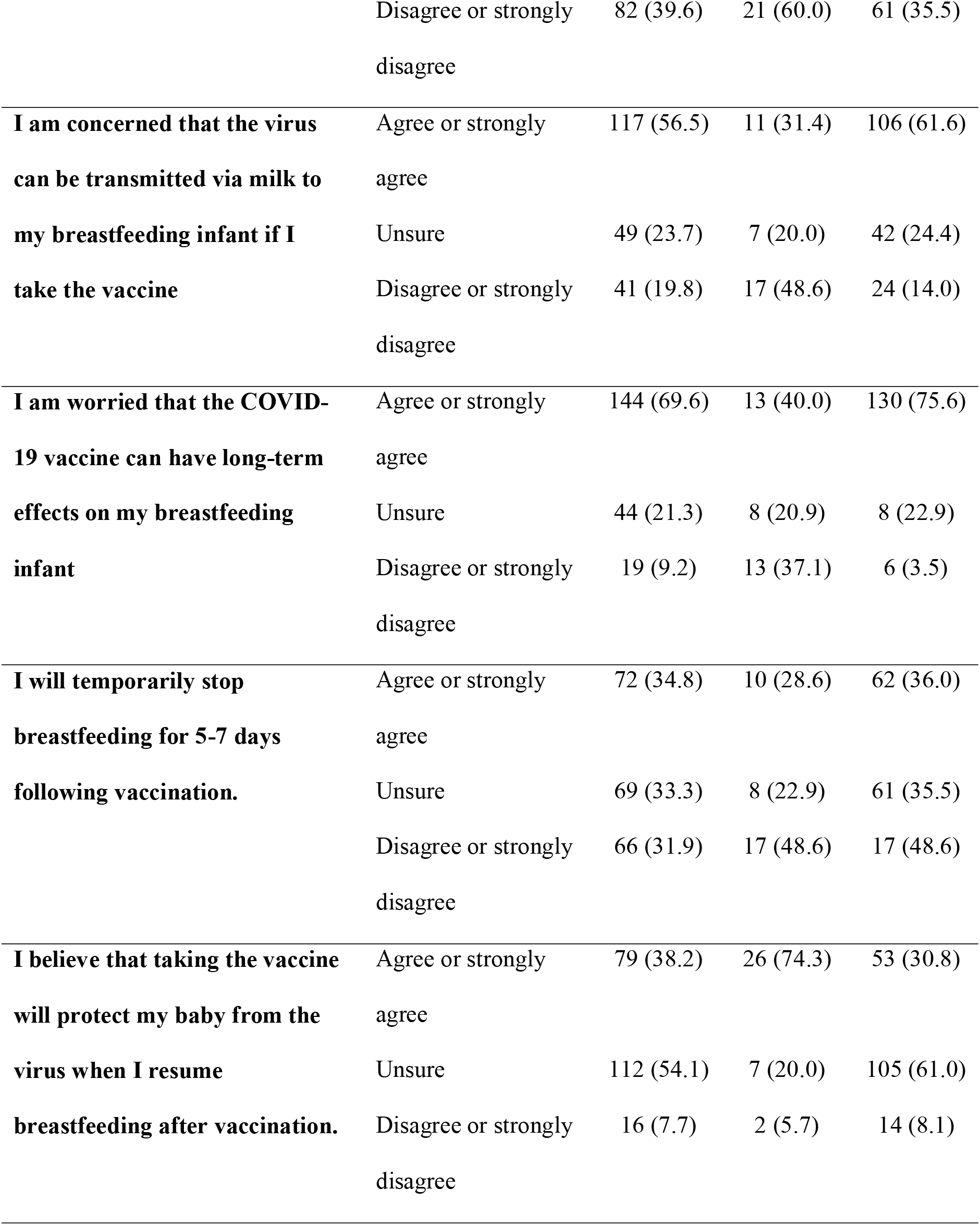
Survey responses for lactating women.

Among lactating women who were unsure or unwilling to take the vaccine (n= 172), many were worried about the side effects that they would experience (147/172, 85.5%), that the virus would be transmitted via breastmilk to the infant (105/172, 61.6%) and anxious about the potential negative long-term effects of the vaccine on the breastfeeding child (130/172, 75.6%). Lactating women also reported to be willing to take the vaccine only if more data on its safety in breastfeeding was available (144/172, 83.7%).

Regression analysis demonstrated that those who received primary school level education were more willing to accept the vaccine than those who had at least an undergraduate degree. Vaccine acceptance was also greater in those with appropriate knowledge regarding vaccine mechanism of action, high maternal perceived risk of COVID-19 and low perceived long-term negative effects on the breastfeeding child (**Table V**).

**Table V.** Multivariate regression analysis for vaccine acceptance in lactating women.

| **Factors** | **Univariate**<br>**Odds ratio (95%**<br>**confidence interval)** | **p** |
| --- | --- | --- |
| **Highest Education level** |  |  |
| No formal education/Primary | Ref | N.A. |
| Secondary/post-secondary | 0.04 (0.01, 0.48) | 0.10 |
| Undergraduate or above | 0.10 (0.01, 0.90) | 0.04 |
| **Knowledge about mechanism of action** |  |  |
| Unsure/Disagree or strongly disagree | Ref | N.A. |
| Agree or strongly agree | 9.44 (1.10, 81.0) | 0.04 |
| **Maternal perceived risk of COVID** |  |  |
| Low or very low | Ref | N.A. |
| Neutral | 0.68 (0.22, 2.06) | 0.49 |
| High or very high | 9.10 (2.40, 34.47) | 0.001 |
| **Perceived long-term negative effects on**<br>**breastfeeding child** |  |  |
| Disagree or strongly disagree | Ref | N.A. |
| Unsure | 0.15 (0.03, 0.33) | 0.01 |
| Agree or strongly agree | 0.08 (0.02, 0.31) | <0.001 |
| **Perceived long-term protection of baby** |  |  |
| Disagree or strongly disagree | Ref | N.A. |
| Unsure | 0.17 (0.02, 1.42) | 0.10 |
| Agree or strongly agree | 0.68 (0.09, 5.01) | 0.71 |

## Discussion

We sought to determine factors associated with COVID-19 vaccine acceptance among pregnant and lactating women in Singapore to guide vaccination efforts in these vulnerable populations. From this survey, we found low vaccine acceptance, varying levels of understanding of vaccine efficacy, and high proportions of safety concerns regarding possible side-effects on the unborn or breastfeeding child in our local population.

Reported rates of COVID-19 vaccine acceptance in pregnant women vary worldwide, from 77% in a study conducted in China to 37% in a study conducted in Turkey.^(20,21)^ Vaccine acceptance in our population appears to be low at 30%, suggesting the need for an action plan to improve acceptance to achieve adequate vaccination rates in this population. Our low vaccine acceptance appears to be guided by a low perceived risk of infection due to low national infection rates, as perceived high COVID-19 infection risk was significantly associated with greater vaccine acceptance in both pregnant and lactating women in our study. The low vaccine acceptance in the survey population may have also been affected by recommendations by the local Health Ministry at the time of the survey. Pregnant women were advised against taking the COVID-19 vaccine, while lactating mothers were discouraged from breastfeeding for 5 to 7 days after vaccination.^(3,4)^ In the United States of America, where organizations support vaccination of pregnant and lactating women, vaccine acceptance appeared to be higher than in our population (44-58% and 55% in pregnant and lactating women respectively).^(16,22)^ Although current recommendations in Singapore have been revised to encourage vaccinations in pregnant and lactating women as on 31^st^ May 2021, it is uncertain whether this will increase vaccine acceptance.

Surprisingly, a larger proportion of pregnant women were agreeable for the vaccine compared to lactating mothers. This was contrary to other studies that reported lower mRNA vaccine acceptance in pregnant compared to non-pregnant or lactating women, which reported concerns regarding potential unknown long-term consequences in the developing fetus among pregnant women.^(15,16)^ A possible explanation is that pregnant women may have been informed of the greater risk for severe disease in infected pregnant women, thus increasing their vaccine acceptance. Pregnant women are also more likely to have had contact with their healthcare providers compared with lactating women, which could have helped to alleviate their concerns and doubts regarding vaccine safety. Nevertheless, this demonstrates that the concerns of both lactating and pregnant women need to be addressed to increase their vaccine acceptance.

Several studies have reported safety concerns of mothers about their child or their own health as the primary reason for low vaccine acceptance amongst pregnant and lactating women.^(16,21,22)^ Similarly, the lack of safety data of the COVID-19 vaccine was a significant factor affecting vaccine acceptability in our population. Pregnant women were concerned about general safety of the vaccine during pregnancy, while lactating mothers were concerned about the possible long-term side effects that the vaccine could have on their child. This is unsurprising, as both pregnant and lactating women were excluded from the initial mRNA-based vaccine trials, limiting the available data on safety and efficacy in these groups. However, since then, observational data from other countries has shown that the mRNA-based vaccine is safe in pregnant and lactating women without any discernible short or medium-term side-effects to the fetus or child.^(23-25)^ Dissemination of this safety and efficacy data in a timely manner is in the key to increasing vaccine uptake in these populations.

While the majority of pregnant women and mothers were aware of the mechanism of action of the mRNA-based vaccines, approximately a third were unsure or unaware of this. Lack of knowledge regarding vaccine mechanism of action was associated with poor vaccine acceptance in both pregnant and lactating women, indicating the potential for educating women on the vaccine’s mechanism of action, so as to increase vaccine acceptance rates. At the time of this survey, only mRNA vaccines were available for vaccination in Singapore. However, as other types of vaccines become available in the future, adequate public education regarding the different mechanism of actions of each vaccine may become important.

We found that lower monthly household income and lower education level were associated with higher vaccine acceptance in pregnant and lactating mothers respectively. This is contrary to other studies where higher education level and socioeconomic status were associated with greater vaccine acceptance.^(26, 27)^ A possible explanation for this is that women who were better educated and of higher socioeconomic status were better informed about the lack of safety data on the vaccine. This may also explain our finding of higher vaccine acceptance in pregnant compared to lactating women, as a smaller proportion of pregnant women had an undergraduate degree or higher (44.3% vs. 65.7%). Indeed, we found that majority of both pregnant and lactating women expressed willingness to take the vaccine when more safety data became available.

A limitation of our study is the cross-sectional nature and the short study period of 3 months. Since conducting our survey, government recommendations regarding vaccination in these groups of women have evolved, with an emphasis that it is safe and recommended. As such, perceptions of pregnant and lactating women may have also changed. However, we believe that our results are still valid in informing educational strategies in women who are unsure or unwilling to take the vaccine despite changes in recommendations. In addition, we measured vaccine acceptance via participant report, and not actual vaccination rates. Reported intent may not translate into actual behavior on vaccination,^(28)^ and whether addressing safety concerns would be adequate in increasing vaccine uptake remains to be determined.

To our knowledge, this is the first survey on COVID-19 vaccine perceptions in pregnant and lactating women in Singapore. Our study is timely in informing education programs and vaccination efforts in these groups to design a targeted action plan. Based on our results, an educational brochure is currently being prepared to better inform pregnant women about COVID-19 vaccination. We also propose the use of other platforms such as mass media and social media to help raise awareness and knowledge to aid improve the acceptance of the COVID-19 vaccine. With the expansion of the local vaccination program to pregnant and lactating women, it is important for educational messages to be individualized such that concerns regarding safety and efficacy are adequately addressed.

## Conclusion

COVID-19 vaccine acceptance in pregnant and lactating women in Singapore is generally low. Factors affecting low vaccine acceptability among both pregnant and lactating women include a low perceived infection risk, and concerns of unknown safety for the mother and child. Addressing safety concerns and infection risk may help improve COVID-19 vaccine acceptance in pregnant and lactating women in Singapore.

## Data Availability

data available

